# Implementation Research to increase coverage of pneumonia treatment in under-five-year-old children in a North Indian district: Study Protocol

**DOI:** 10.1101/2025.03.26.25324663

**Authors:** Barsha Gadapani Pathak, Yasir Bin Nisar, Tarun Madhur, Naveen Garg, Shamim Ahmad Qazi, Sarmila Mazumder

## Abstract

**Introduction:** The National Family Health Survey-5 has reported an under-five mortality rate of 41.9 per 1000 live births in India. Pneumonia, one of the leading causes of under-five mortality contributes substantially to this figure. The Indian government has made efforts through multiple national programs, but pneumonia-specific mortality remains high. The Government of India revised their Childhood Pneumonia Management Guidelines in 2019 to improve under-five pneumonia prevention and management. This study aims to achieve a high population-based coverage of pneumonia treatment for under five-year-old children in the Palwal district of India.

**Method and analysis:** This research utilizes a quasi-experimental pre-post design and a mixed-method approach. The study is in Palwal district, which has a population of about 1.3 million and numerous government healthcare facilities. The catchment area focuses on Health and Wellness Centers, which are the most accessible public health facilities for the community, located close to the homes.The intervention focuses on early pneumonia identification by community healthcare workers and referral of these cases, prompt care-seeking by caregivers from appropriate healthcare providers, and appropriate diagnosis, management of cases at out-patient basis by healthcare providers and referral of severe cases to higher facilities.. The research team (RT) will provide technical inputs and guidance to government staff, who will implement the interventions. The RT comprises three teams: the implementation support team, the program learning team, and the outcome measurement team. The study has three phases: I-Formative research, II-Model optimization, and III-Scale-up and concurrent evaluation. Data will be collected using electronic tablets and the REDCap platform. Data analysis will be performed using Stata 16 and NVivo.

**Ethics and dissemination:** Ethical approval was granted by the ethical committees of the Society for Applied Studies (**SAS/ERC/IR Pneumonia/2021**) and the World Health Organisation (**WHO/ ERC.0003652**). This research is registered in the clinical trial registry **CTRI/2021/03/031622**. [Registered on: 01/03/2021]. Dissemination meetings in the study country will share results with stakeholders, including Ministry of Health officials, health managers, families of under-five children, community leaders, and academia, to discuss national health program implications. Results will also be shared regionally and globally, with publications and presentations encouraged in national and international forums.

**Strengths and limitations of this study:**

- Provides empirical evidence on implementing a national guideline on childhood pneumonia management in a resource-limited district, highlighting real-world challenges and solutions.
- May demonstrate the potential of community-level health workers and primary care healthcare providers in increasing the coverage of pneumonia treatment among children, emphasizing the importance of localized, context-specific strategies.
- Integration of key outcome indicators into the Health Management Information System (HMIS) may enhance public health data systems and ensure sustainability in management.
- May contribute to the global discourse on under-five pneumonia management in low- and middle-income countries, providing insights that can be adapted to similar settings globally.
- However, Reliance on self-reported data from caretakers may introduce a potential recall bias.

Additionally, a broad operational definition of pneumonia may lead to false-positive cases reported to healthcare facilities.

## BACKGROUND AND RATIONALE

The National Family Health Survey-5 (NFHS-5), conducted from 2019 to 2021, reported an under-five mortality rate of 41.9 per 1000 live births in India. (1) Data from national surveys between 2016 and 2018 revealed that the top three causes of under-five deaths in India were lower respiratory infections (17.9%), neonatal preterm birth (15.6%), and other neonatal disorders (14.3%). (2) Among infectious diseases, pneumonia is the primary contributor to under-five mortality, accounting for 15%. (3) There are an estimated 30 million annual cases of acute respiratory illness (ARI), including 3 million cases of severe pneumonia. These statistics highlight the critical public health concern posed by under-five pneumonia and its substantial impact on India’s goal to reduce under-five mortality to 23 per 1000 live births by 2025. (4)

The Indian government has consistently supported pneumonia control measures through multiple national programs, Home-based Newborn care (HBNC), Home-based young child care (HBYC), Integrated management of neonatal and childhood illness (IMNCI), Facility-based integrated management and neonatal and childhood illness (F-IMNCI), Poshan Abhiyan, Mother’s absolute affection (MAA) program, Universal Immunization Program/Mission Indra Dhanush, Vitamin A supplementation, Infant and Young Child feeding program, Facility based newborn care (FBNC), to address childhood infections prevention and enhance under-five infection management. (4) Despite these efforts, India still witnesses an annual pneumonia-specific mortality of 0.14 million. (4) Only 22% of caretakers seek appropriate care for acute respiratory infections due to factors like limited awareness, financial constraints, healthcare accessibility issues, reliance on home remedies, and insufficient community involvement. (5) Front-line health workers also face challenges, lacking the necessary skills and resources. (6) The primary challenge lies in the effective implementation of these well-intentioned national guidelines and programs. (4)

Considering the substantial burden and the pressing need to intensify under-five pneumonia prevention and management efforts, the Government of India (GoI) launched the Childhood Pneumonia Management Guidelines in 2019. (4) In pursuit of ensuring the program’s visibility and long-term sustainability, the Social Awareness and Actions to Neutralize Pneumonia Successfully (SAANS) campaign was initiated, aligning with the Protect, Prevent, and Treat (PPT) interventions for childhood pneumonia. (4, 7) The primary objective of this campaign is to significantly expand the reach of pneumonia awareness and management which may eventually reduce the pneumonia burden. (4) The strategic vision revolves around the utilization of health and wellness centers (HWCs), established under the Ayushman Bharat Yojana, as central nodes for early identification, treatment, and timely referral of severe cases of under-five childhood pneumonia to tertiary healthcare facilities for life-saving interventions. (4) Nevertheless, to implement the current Government of India (GoI) pneumonia management guidelines, certain significant requisites may be the comprehensive enhancement of healthcare worker training and skill development, the augmentation of hospital infrastructure, and the active engagement of local communities. (8-10)

Considering the above context, we propose to conduct this implementation research aiming to achieve high population-based coverage of the GoI childhood pneumonia management guidelines, defined as 80% or more under-five children in the selected study area, receiving the full course of the pneumonia treatment. (4) All the activities will be conducted with continuous government engagement.

Furthermore, within the framework of this project, we aim to augment all supplementary programs contributing to pneumonia management. This also includes the strengthening of Health and Wellness Centers (HWCs) and the enhancement of pneumonia management across various government healthcare facilities, encompassing primary health centers, community health centers, and district hospitals. We expect that synthesizing the operational lessons and delivery strategy insights from this implementation research will facilitate the scaling up of pneumonia treatment and case management in similar settings, with necessary contextual adaptations.

### Study Aims and Objectives

This study aims to develop an optimized, sustainable and scalable model to achieve high population-based coverage of pneumonia management among under-five children.

#### Primary objective

To achieve high population-based coverage, defined as full treatment of pneumonia in 80% or more under-five children in the defined study area, as recommended by the GOI guidelines. (4)

#### Secondary objectives

i) The evaluation of recovery proportion among children following seven days of treatment, ii) identification of treatment failure proportion, iii) assessment of compliance with pneumonia treatment, iv) proportion of children who complied with hospital referral as recommended in the guidelines, v) duration and details of in-patient treatment, vi) exploration of the knowledge, perception, and experiences of healthcare workers, community health workers, and beneficiaries regarding pneumonia management implementation and vii) promote pneumonia treatment among girl children in the context of to “Save the Girl Child” program.

## METHODS AND ANALYSIS

### Study design

This implementation research includes a mixed-method approach aiming to scale up the management of under-five pneumonia and utilizes a quasi-experimental pre-post design. Our approach involves a non-sequential, repeated, cyclical, and iterative implementation process, characterized by a series of overlapping cycles that involves implementation, process learning with both quantitative and qualitative feedback, and ongoing quantitative coverage evaluation. (11) Regular meetings in collaboration with government officials, will provide a platform for deliberation on the implementation progress, the insights acquired, the assessment of model performance, and the continuous enhancement of the implementation process. (11)

### Implementation science framework

we will employ a flexible and adaptive strategy, actively engaging relevant stakeholders, thoroughly examining contextual factors, identifying implementation barriers and facilitators, allocating essential resources as needed, and establishing a continuous monitoring system. These components collectively contribute to our aim of ensuring the success and sustainability of this guideline, embodying key elements of a robust implementation framework. (11)

### Operational definitions

In this, pneumonia is defined as the mother/caregiver reporting any episode of cough along with either fast breathing or difficult breathing or stridor (“khar khar” or “khad khad”) or chest indrawing or pneumonia (“pasliyon ka chalna”) or any danger signs (like axilliary temperature < 35.5 or >=37.5 ^L^C or not able to feed or convulsion or movement only when stimulated or no movements at all among child less than 59 days and inability to breastfeed or drink or vomits everything or lethargy or reduced level of unconsciousness or convulsions among children aging 2 – 59 months of age) in the last 28 days.(4) In a low-resource setting like India grassroots-level workers like Accredited social activists (ASHA) and caregivers, sometimes non-literate, are the ones reporting cases of pneumonia to the health facility. In this context, it is challenging to employ a definition of pneumonia that requires access to laboratory or radiological examinations. Considering the existing capacity of the health services, we need to employ a definition that is based on clinical signs that can be recognized by caretakers or ASHAs.(5) Hence, the definition of under-five pneumonia, in this study, is broad (with low specificity) for operational reasons. Additionally, this operational definition has been employed in other studies which were conducted in similar settings in India. (5)

### Study setting

The study will be conducted in the Palwal district, located in the northern Indian state of Haryana, and covers an area of 1,359 square kilometers, including three administrative divisions known as tehsils: Palwal, Hodal, and Hathin.(12) It comprises 282 villages, 237 Gram Panchayats, 1 municipal council, 2 municipal committees, 3 sub-divisions, and 4 development blocks, and is located approximately 80 kilometers from the capital of India. This district serves a population of 1273665 through 1 District Hospital (DH), seven Community Health Centers (CHCs), 26 Primary Health Centers (PHCs), 100 sub-centers, 49 functional health and wellness centers, and 1108 Anganwadi centers. The PHCs target to serve around 49000 people each with 10 SCs catering to about 12000 to 13000 people each. (12)The district has 37 medical officers at CHCs and PHCs, 176 medical officers, and other healthcare professionals at the district hospital. The average literacy rate is 69.32%, and 12% of the population is under five years of age with an annual birth rate of 25 per 1000 population. (12, 13)

The catchment area for our implementation research will be the HWCs of Palwal district. The HWCs were established under the Aayushman Bharat – Haryana Health Protection Mission (AB-HHPM), and currently, 49 are functional in the Palwal district to provide comprehensive primary health care services in proximity to residents’ homes. (13)

The district health authorities will determine the number of HWCs to be assessed from the 49 functional centers and subsequently share the list with the research team. After the preliminary assessment, the research team will select ten HWCs for inclusion in the study. We will exclude HWCs that lack appropriate infrastructure and human resources because these are basic prerequisites to implement the program. It would be time-consuming to put these into place and may not be feasible within the study period. Furthermore, in addition to the selected HWCs, the research team will conduct other facilities assessments (PHCs, CHCs, and DH) that cater to the population referred from these HWCs. The catchment area of this study will have an estimated population of 1,07,440, consisting of 42 villages and 9897 under-five populations. (12)

**Fig 1:**
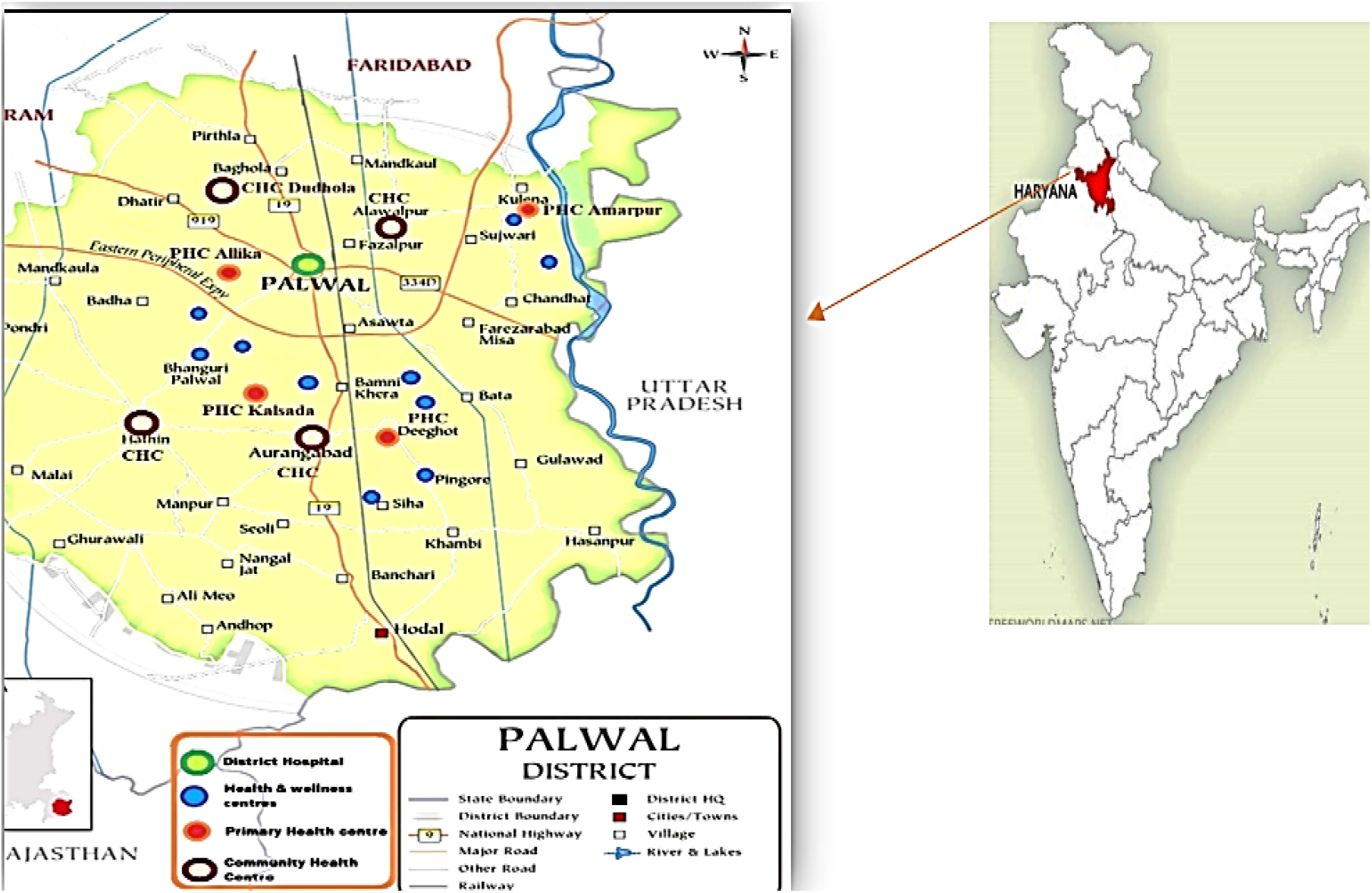
Map of the study area.

#### Eligibility and inclusion criteria

The mothers/primary caregivers of the under-five children, healthcare workers, in the facility and community of the catchment area will be included in this study.

### Sample size

A recent study in the same district found that the prevalence of pneumonia among under-five children in the last two weeks was around 5%.(14) Currently, approximately 25% of under-five children seek appropriate care for acute respiratory illness, indicating a program coverage of around 25%.(5, 15) With the introduction of the new implementation strategy, we expect an increase in pneumonia treatment coverage by 80%. To account for potential non-response and to estimate 80% coverage of appropriate treatment at baseline and end-line surveys for under-five children with pneumonia cases (as per the operational definition) a conservative sample size of 120 is to be included. Additionally, based on these assumptions, the estimated sample size at a 5% level of significance for each survey (baseline and end-line) and a varied range of coverage is shown in **Figure 2**.

**Figure 2:**
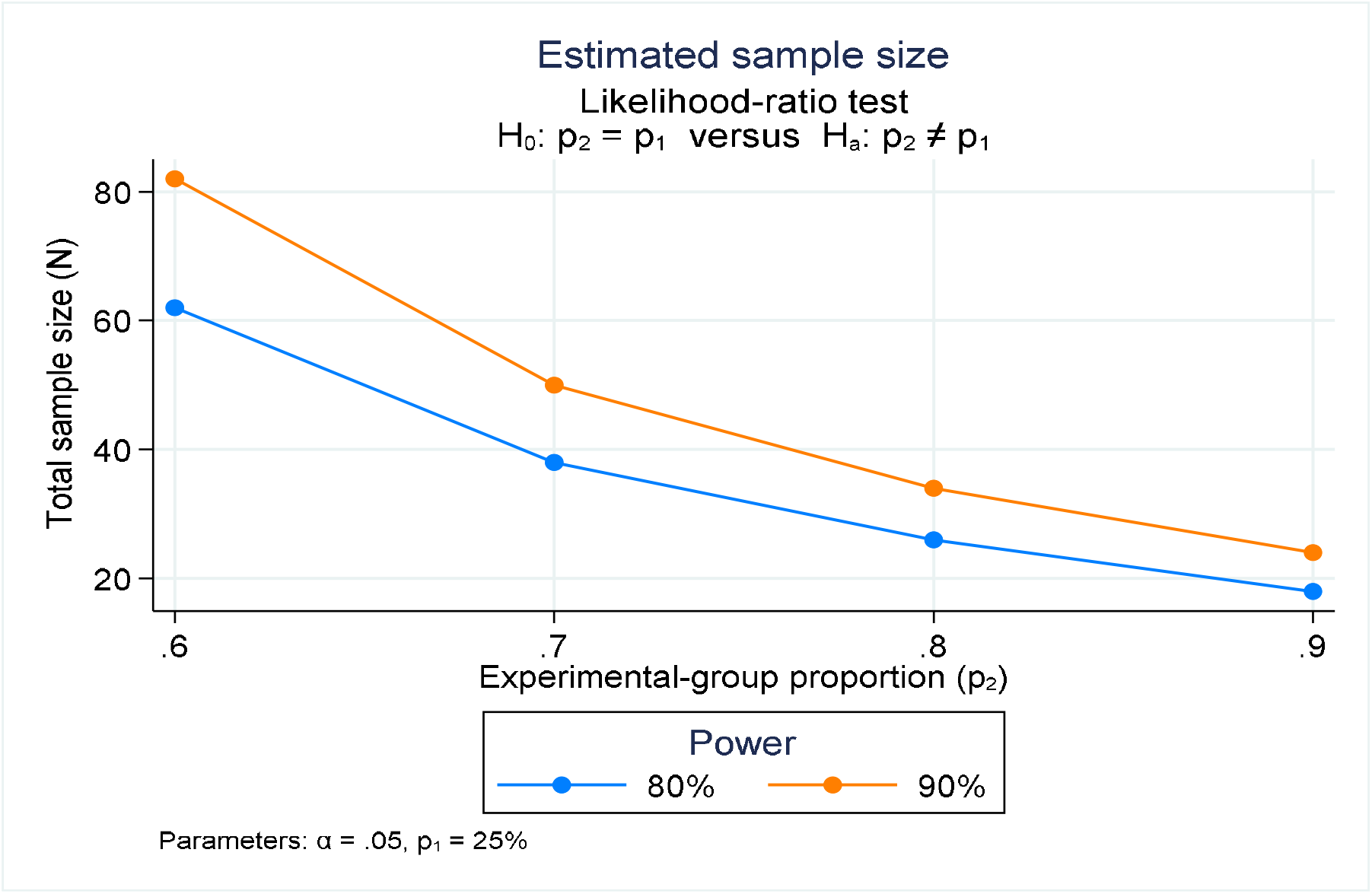
Sample size estimation.

The total number of children to be surveyed is calculated based on a prevalence rate of 5%, resulting in an estimated sample size of 2,400. (5) Considering the proportion of 5-year-olds in the population (12% based on census data) and the average household size (4.8 as per the NFHS 5 report for Haryana), the number of households to be covered is calculated as 4,167 households. (16). These households will be selected using Probability Proportional to Size (PPS) sampling methods from the defined study area served by the 10 HWCs (17) Since community-level healthcare workers maintain a list of under-five children in each household, systematic random sampling will be used to select children within the chosen households.

### Outcomes

The primary outcome of the study is to find the coverage of pneumonia management from appropriate sources. The secondary outcomes are total recovered cases after seven days of treatment, treatment failure, compliance to management as recommended by healthcare providers, in-patient cases duration and treatment provided, and gender-wise distribution of diagnosed, treated, and complaint cases.

### Intervention package

The intervention package will be aligned with the GoI guidelines which include early identification of pneumonia cases through Accredited Social Health Activists (ASHAs) and empowering families, prompt care-seeking from appropriate providers for pneumonia treatment, and prompt referral of severe pneumonia cases with pre-referral treatment. At the outpatient department (OPD) of facilities (HWCs/PHCs/CHCs/DH), treatment providers need to be available, and competent and have access to appropriate logistics (equipment, regular supplies of medicines). Pulse oximeters will be provided at the OPDs of these facilities to assess oxygen saturation for improved assessment/evaluation of severe cases requiring referral.

For children, (0-59 days) with Possible serious bacterial infections (PSBI) a prereferral dose of gentamycin and amoxicillin will be provided by the health workers at the HWCs and then cases are referred to higher facilities. But if a referral is not possible/denied the package also aims to provide treatment to children aged 0 to 59 days in the outpatient department of primary-level care. In cases where caregivers are unable to bring their infants to the health facilities for the full course of injection gentamycin and oral amoxicillin for 7 days will be provided by the ANM to cases (0-59 days) at home. (4, 18, 19)

In discussions with the government, strategies will be finalized to promote treatment in female children, such as providing additional incentives to ASHAs for referring or accompanying girl children for pneumonia treatment, free transportation for girl children to facilitate referral for out-patient treatment and providing free food or compensating daily wages lost due to hospital admissions.

At the post-facility level, ASHAs will be rewarded for ensuring full compliance with the treatment course in female children. The decision to address the appropriate treatment of pneumonia in the private sector, where a large number of children with pneumonia seek care, by involving private providers in the discussion with the government partners, will be based on the formative research findings. Appropriate use of bronchodilators in children with wheeze at health facilities and identification of conditions mimicking pneumonia for rational therapy will also be included. Finally, recognition of cases that do not have pneumonia and do not require antibiotics but may benefit from supportive treatment.

In the process, all collateral programs supporting pneumonia management will be strengthened like Janani Shishu Suraksha Karayakram (for free transportation, medicines, and diagnostics test availability), Comprehensive Primary Health Care (strengthening care at HWCs), Home-based Neonatal Care (improving the quality of care by ASHAs), etc. All the activities will be conducted with deep government engagement.

**Fig 2.**
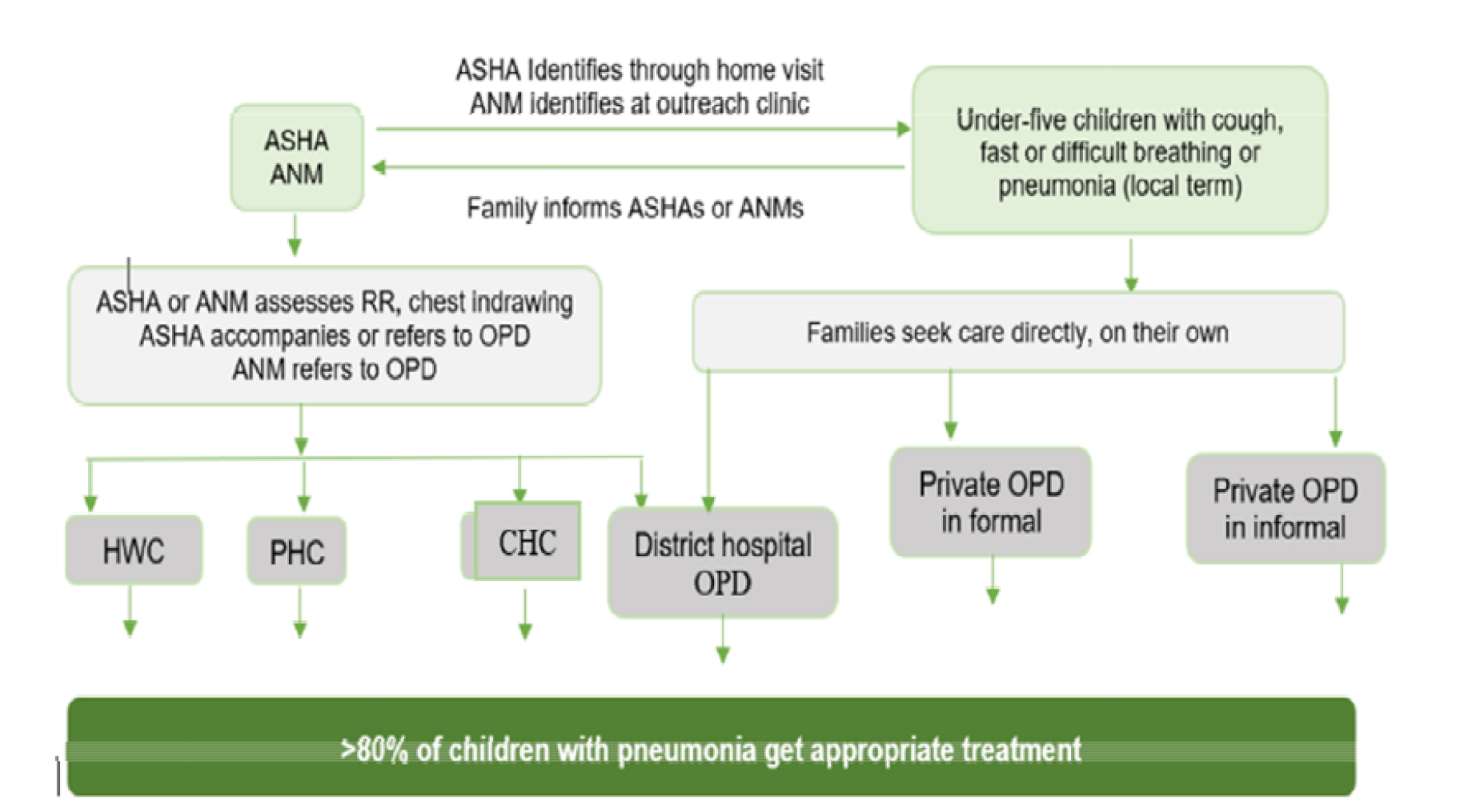
The conceptual framework for increasing coverage of pneumonia treatment.

### Research teams

There will be three teams: i) the Implementation Support Team (IST), ii) the Program Learning Team (PLT), and iii) the Outcome Measurement Team (OMT).

The IST will provide technical support to government partners and assist in program implementation. They will facilitate government staff training in the study area, identify process gaps for refinement, and develop tools and job aids. Additionally, the IST will support community awareness activities and data collection from ASHA records and healthcare facilities.

The PLT, composed of qualitative researchers, will assess fidelity and compliance through methods like focus group discussions, in-depth interviews, and observations. They will provide prompt feedback to the IST and deliver weekly reports.

The OMT will focus on measuring primary and secondary outcomes. They will conduct baseline and end-line cross-sectional surveys in randomly selected households within the study area, assessing population-based program coverage using a randomized sample selection method.

### Implementation strategy

The implementation plan for promoting the treatment of pneumonia in under-five children will follow a systematic approach involving formative research, engagement of key stakeholders, model development, implementation, and evaluation. (Fig.3) The study will consider the GoI childhood pneumonia guidelines and develop model 0 through expert consensus. In-depth interviews and focus group discussions will be conducted to gain a comprehensive understanding of stakeholders’ perspectives, knowledge, and practices related to pneumonia management. For the implementation of guidelines, the key stakeholders, including mothers/primary caregivers, health care workers, community health workers, and government partners, will be identified and actively engaged in the study catchment area.

The formative research findings will be analyzed to identify pneumonia management barriers and facilitators, and a Model 0+ implementation strategy will be developed. This model will include strategies to create caregiver awareness, encourage care-seeking at appropriate health facilities, improve community health workers’ skills in case identification and referrals, and provide training and capacity building for other healthcare workers to enhance early identification and treatment of pneumonia cases. Workshops will be conducted with the research team and government partners to refine and finalize Model 0+ for local alignment and needs.

Following the development of Model 0+ will be implemented in a learning block comprising five randomly selected health and wellness centers, with the close collaboration of government partners, to provide initial handholding and support for implementation. Monitoring and evaluation of process indicators such as access, utilization, compliance, adherence, and fidelity to childhood pneumonia management guidelines, as well as health facility components like infrastructure, human resources, delivery, availability of medicines and supplies, and documentation, will be conducted. Evaluation data will be analyzed and shared with the government implementation team monthly to guide decision-making. Any inconsistencies identified between the evaluation data and existing system data will be addressed through corrective actions.

The research findings from the learning block will be used to enhance Model 0+, leading to the development of Model 1. Subsequently, a series of rapid cycles involving concurrent implementation, monitoring, and strategy refinement will be conducted in the remaining five HWCs. These iterations will take place in multiple co-design workshops, with significant involvement from government authorities. The iterative process will continue until an optimized and comprehensive strategy/model is achieved with the potential for achieving high coverage and quality of pneumonia management among under-five children in the study areas. (Fig 3)

**Fig. 3.**
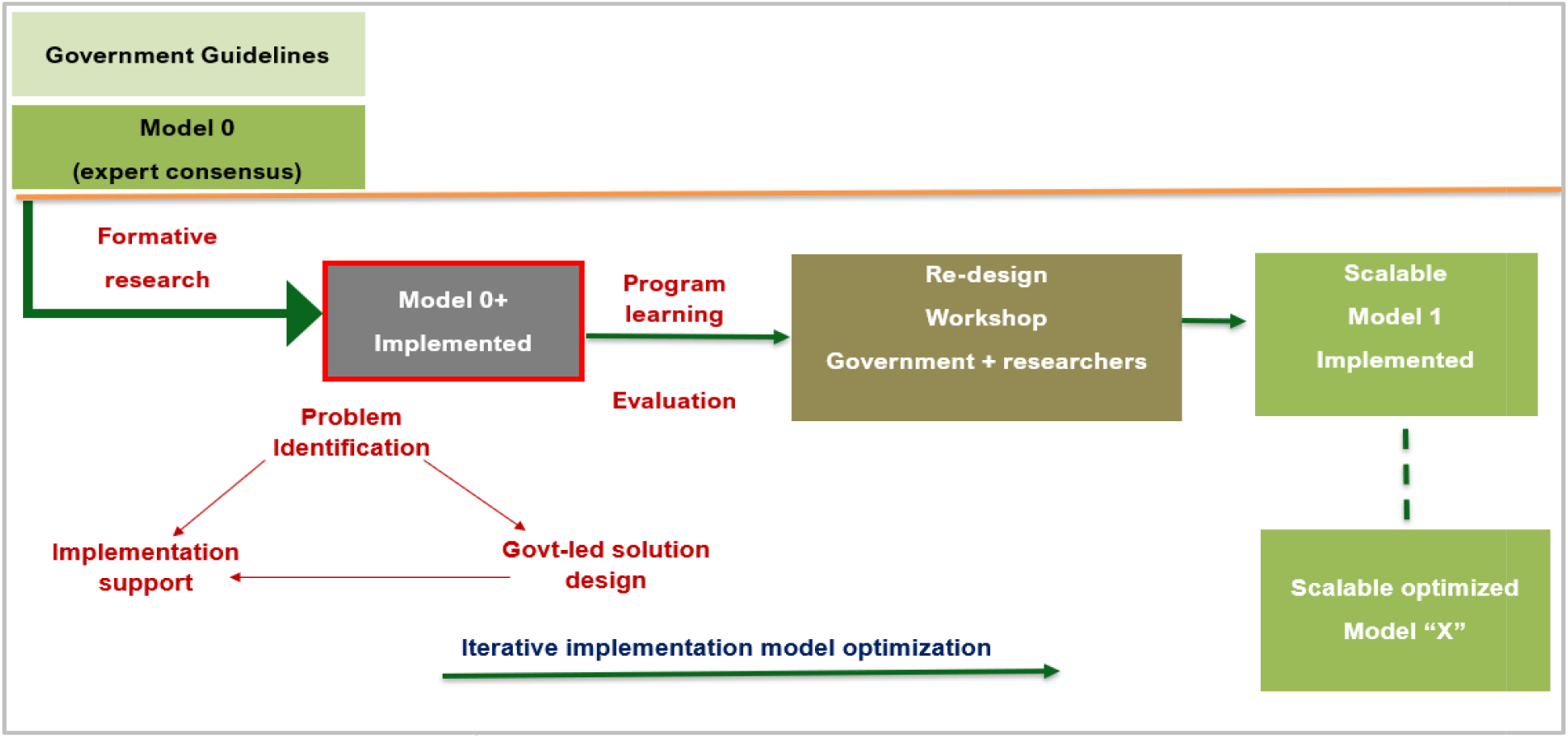
Iterative cycles of model optimization. Figure adapted from World Health Organisation.

### Data Management and data analysis

#### Quantitative Data

The study’s data management will be handled by a data management center (DMC) located in the study field office. The DMC will be using the REDCap (Research Electronic Data Capture) which is a secure web application for building and managing clinical research data. It is a powerful and flexible tool for data management that can help streamline the research process and improve data quality. The quantitative data will be managed and reviewed by the respective team coordinators, and the data entry system will have inbuilt range and consistency checks. Cross-sectional survey data will be collected electronically and transferred in real-time to a local server at the DMC, where checks across forms and logical error checks will be run. Queries will be generated for the study team’s resolution, corrections will be incorporated into the cleaned data set, and the tables for quality assurance will be generated at the Society for Applied Studies (SAS) office in Delhi daily.

Four types of data will be collected, i) health systems data, ii) routine patient data, iii) cross-sectional household survey data, and iv) information on knowledge of pneumonia signs and symptoms and its management. Descriptive and inferential statistical analyses will be performed, including simple proportions, and generalized linear models (GLM), using the univariate and multivariate regression analyses. The study will determine the prevalence of pneumonia, overall coverage of pneumonia treatment, compliance to treatment, and treatment failure, and identify predictors of treatment failure.

### Qualitative data

Qualitative data will be collected to understand the experiences of healthcare providers and community health workers in treating pneumonia, as well as the perceptions of mothers and family members. Field notes will be recorded and transcribed from FGDs and IDIs. We will use the N-vivo software package for data management and analysis. Data analysis will be conducted simultaneously with data collection, and relevant text will be coded daily. Codes will be descriptive or analytical, based on the research question, and grouped into themes and sub-themes. A framework analysis will be used, primarily a case and theme-based analysis. A matrix will display information to examine information across rows and down columns for theme development and to maintain context. Findings will be weighted by identifying key themes and estimating the number of times they appeared and the number of respondents who mentioned them.

## ETHICAL CONSIDERATIONS

The study will adhere to Good Clinical Practice guidelines and obtain ethical clearance from the site ethical review committee and World Health Organization, as well as any necessary national, regional, or state approvals. Before enrolment, the parent or caregiver of each participant will be provided with a written informed consent form in the local language, and non-literate parents will have the form read to them with a thumb imprint obtained. The healthcare workers involved in the in-depth interviews and focus group discussion will also be provided with a written informed consent form. This protocol complies with the reporting guidelines of the STROBE (Strengthening the Reporting of Observational Studies in Epidemiology), and the Standard for Reporting Qualitative Research (SRQR) guidelines. (20, 21)

### Confidentiality and data handling

Data custody will be with the data coordinator, who will grant access to the data manager. All computers and screens will be password protected and staff will receive user access authorization and ICH-GCP training to ensure confidentiality and data protection. Health information will be computerized in India and collected electronically using tablets. The real-time data will be transferred and securely stored on a cloud-based server, accessible only to authorized personnel of the study. Personal information and identifiers will be kept confidential in a secured location accessible only to Indian investigators, and regular backups will be taken. De-identified data will be shared with WHO for further analysis, with identifiable information such as names, addresses, and contact numbers removed. Participants cannot be identified from the data directly or indirectly.

## PATIENT AND PUBLIC INVOLVEMENT

Before the development of this protocol, we had a pilot phase which had extensive engagement within the research team, government stakeholders, community representatives and beneficiaries like the caregivers of under-five children and their families.

## DISSEMINATION OF FINDINGS

Dissemination meetings will be organized in the study country to share the results and discuss their implications for the national health program. Stakeholders including the Ministry of Health program managers, district and sub-district health managers, community leaders or representatives, and academia will be invited to attend. Results will also be disseminated at the regional and global levels. Publication in national journals and presentations in national and international meetings will be encouraged.

## DISCUSSION

Pneumonia remains the leading infectious disease-related cause of death among children under five in India, responsible for 15% of such fatalities. Achieving the National Health Policy’s goal of reducing under-five mortality to 23 per 1000 live births by 2025 necessitates a substantial reduction in pneumonia-related mortality to 3/1000 live births. This implementation research strives to develop a scalable strategy for pneumonia management, focusing on extensive population-based coverage of under-five pneumonia treatment recommended by the Government of India guidelines. The study will comprise formative research, model optimization, and evaluation across the selected study area in Palwal district, Haryana, India, encompassing outpatient and inpatient pneumonia management.

The study’s optimized pneumonia management strategies, developed following implementation research principles, can be applied in similar low- and middle-income countries, enhancing coverage and reducing under-five pneumonia case fatality rates. Integrating key outcome indicators into the Health Management Information System (HMIS) promotes accountability and sustainable scale-up. Furthermore, the research identifies system-level factors impacting healthcare system performance, allowing targeted improvements in areas like training, supply chain, and healthcare worker payments, thereby strengthening overall coverage of pneumonia treatment.

In Indian and other low to middle-income settings, cluster-randomized controlled effectiveness trials have explored the management of uncomplicated pneumonia in infants aged 7 days to 59 months, employing community-level health workers and it reported that trained community-level workers like Accredited Social Health Activists (ASHAs) can effectively manage the cases and are generally accepted in this role by the community. (22) However, the Indian government chose not to employ this strategy due to concerns about ASHAs lacking the essential skills and educational qualifications for managing under-five illness. Consequently, the Government of India’s guidelines focus on ASHAs primarily identifying pneumonia cases and referring them to the nearest Health and Wellness Centers (HWCs) for treatment. This implementation research aims to assess the practicality of implementing the guidelines and explore alternative strategies in collaboration with the government.

The proposed study exhibits multiple strengths, such as its concentration on a resource-constrained environment and its adherence to the core principles of implementation research. This entails addressing practical obstacles, formulating strategies to surmount these challenges, and creating an optimized implementable model derived from acquired knowledge and insights. This model can subsequently be adapted to similar settings, allowing for broader scalability while considering contextual variations.

However, we must acknowledge that the study findings may be limited by the reliance on self-reported data from caretakers, which may be subject to recall bias. This study uses a broad operational definition of pneumonia that can be recognized by caretakers and community health workers, which may lead to an increase in false-positive cases reported to healthcare facilities. While the current study primarily concentrates on expanding the adoption of recommended interventions outlined in the guidelines, it is imperative to investigate the impact of the range of interventions as specified in the guidelines. This could be achieved through a hybrid type-II design, which falls beyond the purview of our present study. (23) To achieve the desired power for outcomes, a large sample size will be required, which mandates extensive funding and resources.

## CONCLUSION

In conclusion, this study underscores the importance of addressing healthcare-seeking behavior among caretakers, improving the capacity of healthcare providers, and increasing coverage of appropriate management to reduce pneumonia-related mortality in under-five children.

Implementation research can help identify system-level factors that may impact the performance of the healthcare system in managing pneumonia cases and contribute to strengthening the capacity and effectiveness of the healthcare system in treating pneumonia.

## Data Availability

The data is available on resonable request to the corresponding author Dr Sarmila Mazumder and/or first author Dr Barsha Gadapani Pathak

## List of abbreviations

GoI: Government of India
ASHA: Accredited social health activists
ANM: Auxilliary nurse midwives
HWCs: Health and wellness centers
PHC: Primary health care centers
CHC: Community health centers
SAANS: Social awareness and actions to neutralize pneumonia

## Declarations

### The ethics approval and consent to participate

Ethical approval was granted by the ethical committees of the Society for Applied Studies (SAS/ERC/IR Pneumonia/2021) and the World Health Organisation (WHO/ ERC.0003652). Consent will be obtained from healthcare and community workers after reading out the information sheet to them. The participants can withdraw or request to stop recording the interviews at any time.

### Consent for publication

Not applicable.

### Availability of data and materials

Not applicable.

### Disclaimer

The authors alone are responsible for the views expressed in this article and they do not necessarily represent the views, decisions, or policies of the institutions with which they are affiliated. YBN is a staff member of the World Health Organization. The expressed views and opinions do not necessarily express the policies of the World Health Organization.

## Author contributions

SM and BGP conceived the study. SM, BGP, TM, NG, NB, YN, and SAQ were involved in reviewing the articles and subsequent drafts of manuscripts. All authors read, critically reviewed, and approved the final manuscript.

## Acknowledgements

We would extend our gratitude to the National Health Mission, Haryana, and the Chief Medical Officer of District Hospital, Palwal, for their extensive cooperation during the initial discussion of this research. We are thankful to the entire study team and specifically senior coordinator Mr. Mandeep Singh, who was instrumental in initiating the discussions of research-related activities with the government.

